# Therapist-Delivered Video CBT for Hoarding Disorder: A Retrospective Observational Study of Clinical Outcomes from a Large Real-World Sample of Adults

**DOI:** 10.64898/2026.05.14.26353262

**Authors:** Jamie D. Feusner, Clare C. Beatty, Patrick B. McGrath, Nicholas R. Farrell, Mia Nuñez, Nicholas Lume, Larry Trusky, Stephen M. Smith, Andreas Rhode

## Abstract

Hoarding disorder (HD) affects approximately 2–3% of adults and is associated with substantial functional disability and limited access to evidence-based care. The aim of the current analysis was to examine the naturalistic effectiveness of therapist-delivered video cognitive-behavioral therapy (CBT) for HD in a large real-world sample, and to characterize individual-level treatment response, time-to-response, and moderators of outcome.

This retrospective, observational analysis examined clinical data from 305 adults diagnosed with HD who received therapist-delivered video CBT through an online specialty therapy platform between September 2021 and February 2026. Hoarding symptom severity was assessed using the Hoarding Rating Scale–Self Report (HRS-SR). Linear mixed models examined symptom change from baseline to three timepoints: session 10, session 20, and each patient’s final session.

HRS-SR scores decreased from *M* = 22.4 (*SD* = 7.6) at baseline to *M* = 16.4 (*SD* = 8.2) at final session (Hedges’ g = 0.81, 95% CI: 0.68–0.94). By the final session, median percent improvement was 25.0% [IQR: 3.0–46.7%]. A total of 39.3% of patients achieved ≥35% HRS-SR reduction, 27.4% of patients who began above the clinical threshold achieved remission, 36.4% demonstrated reliable improvement, and 22.9% of eligible patients achieved clinically significant change. Among patients who achieved and maintained ≥35% reduction through their final session (n = 120), median time to first response was session 9, with 54.2% responding within 10 sessions. Analyses of secondary outcomes showed significant improvements in clutter severity, depressive and anxiety symptoms, stress, quality of life, and functional disability (Hedges’ g = 0.21–0.47). Greater baseline severity, more sessions, and longer treatment duration significantly moderated outcomes; prior OCD treatment history did not.

Findings suggest that therapist-delivered video CBT for HD, delivered remotely in a real-world setting, produces outcomes consistent with controlled trials and may be a clinically effective and scalable approach for a condition historically underserved by mental health systems.

**Author Summary:** Hoarding disorder is a common and debilitating condition affecting millions of adults, yet most people with hoarding disorder never receive effective treatment. Barriers including cost, limited availability of trained therapists, shame, and reluctance to allow others into the home make access to care particularly challenging. In this study, we examined outcomes for 305 adults with hoarding disorder who received cognitive-behavioral therapy delivered by trained therapists via video through an online specialty platform– the largest real-world study of this treatment approach to date.

We found that treatment produced meaningful reductions in hoarding symptom severity, consistent with what has been observed in controlled clinical trials, and that improvements extended to clutter, mood, quality of life, and everyday functioning. Notably, among patients who responded to treatment, over half showed meaningful improvement within the first 10 sessions, a finding not previously reported for hoarding disorder. These results suggest that video-delivered therapy can reach patients who might not otherwise access care and can produce clinically meaningful outcomes at scale.

Our findings support the continued development and dissemination of video-based treatment for hoarding disorder.

## Introduction

Hoarding disorder (HD) is defined in the DSM-5 as persistent difficulty discarding or parting with possessions regardless of their value, leading to accumulation that clutters living areas and causes clinically significant distress or functional impairment (1–3). It is distinct from ordinary collecting and from hoarding symptoms secondary to other mental or neurological conditions (3,4). Core features include excessive saving, strong urges to acquire, distress around discarding, and severely cluttered living spaces that can no longer be used for their intended purpose (2,3).

Population-based estimates suggest HD affects approximately 2–3% of adults (2,5). Hoarding symptoms typically emerge in adolescence and follow a chronic, progressive course that worsens over decades without intervention (6). HD carries substantial individual burden, with functional disability rates comparable to or exceeding those seen in major depressive disorder, diabetes, and chronic pain across cognitive, mobility, self-care, and social domains (7,8). Beyond individual impairment, HD is associated with serious safety risks, including falls, fires, health code violations, and eviction, and places significant burden on family members and health systems (9–11).

A specialized form of cognitive-behavioral therapy (CBT) is the primary evidence-based treatment for HD. Unlike standard CBT protocols for anxiety or depression, CBT for HD incorporates a distinctive combination of motivational enhancement to address low insight and ambivalence, psychoeducation, cognitive restructuring targeting dysfunctional beliefs about possessions, skills training in organizing and decision-making, and repeated exposure-based practice in sorting, discarding, and resisting acquisition, often conducted directly within the patient’s home (12–14). In research trials, treatment has typically ranged from 16 to 26 sessions, though greater session “dose” does not appear to reliably predict improved outcomes (15,16). Randomized controlled trials support the efficacy of CBT for HD across individual and group formats, with meta-analytic evidence confirming large average symptom reductions (12,15–18). However, clinical results have been modest overall: most participants continue to meet diagnostic criteria for HD at posttreatment, only 24–43% show clinically significant change, and residual symptoms, particularly clutter, remain common (14,15).

Despite these established efficacy data, access to effective care for HD remains severely limited. Many individuals with HD come to the attention of services not through mental health pathways but through housing, social care, or legal channels, often at points of crisis such as eviction or enforced cleaning (19,20). Among those who do seek help, common barriers include cost, limited awareness of treatment options, shame, fear of judgment, and fluctuating motivation (21,22). At the system level, HD frequently falls between service boundaries, with unclear referral pathways and fragmented coordination across mental health, housing, and social care (19,20). Even among those who access CBT, dropout and refusal rates are high, and a substantial proportion of completers do not achieve clinically significant improvement (16,21,23). Together, these barriers create a substantial treatment gap in which the majority of individuals with HD never receive adequate care (14,22).

Telehealth delivery of CBT represents a promising avenue for addressing several of these barriers directly. By removing the need for in-person attendance, remote formats reduce the financial and logistical burden of accessing specialist care, extend reach to individuals in areas with few trained clinicians, and may lower the threshold for engagement among those who experience shame or fear of judgment about allowing others into their home (21,24). Remote formats are also particularly well-suited for HD specifically, as sessions can be conducted directly within patients’ own living environments, enhancing ecological validity and potentially strengthening treatment generalization (24,25). In this way, therapists can guide patients with sorting and discarding virtually in their home without time-consuming travel or needing patients to bring boxes into their office. They can also see clutter in the patients’ homes to help monitor progress. In a pilot trial of video teleconferencing-delivered group CBT, approximately 32% symptom reduction was achieved with low dropout and 30% full recovery at six-month follow-up in adults with HD (26); webcam-delivered individual CBT and blended formats combining video sessions with between-session internet support have similarly demonstrated feasibility and preliminary evidence of symptom improvement (27–29). However, existing studies are largely limited to small pilot samples with few controlled comparisons, limiting conclusions about efficacy at scale (24,30). No study to date has examined therapist-delivered video CBT for HD in a large, real-world clinical sample.

The present study addresses this gap by examining real-world treatment outcomes for adults with HD who received therapist-delivered video CBT through a specialty online platform. Treatment was delivered by therapists trained in CBT for hoarding via individual video sessions with supplemental between-session support including therapist messaging and an online peer community. This retrospective analysis of naturalistic clinical data included patients who completed varying numbers of sessions, over varying treatment durations, and at varying intensities. The primary aim was to examine changes in self-rated hoarding symptom severity from baseline to sessions 10, 20, and each patient’s final session. Secondary aims examined whether treatment was associated with improvements in photograph-based clutter severity, depression, anxiety, stress, functional disability, and quality of life. Additional aims examined individual-level treatment response and remission rates across a range of thresholds, session-level time-to-response among patients who achieved clinically meaningful improvement, and potential moderators of treatment outcome including baseline severity, treatment dose, age, sex, comorbidity status, and prior OCD treatment history. To our knowledge, this represents the largest naturalistic study of therapist-delivered video CBT for HD to date.

## Results

### Sample

The final analytic sample included 305 adults with HD (*M*age = 45.6 years, *SD* = 14.0; 75.9% female; 60.3% White). Comorbid diagnoses were common, with 51.1% having a comorbid anxiety disorder and 50.5% a mood disorder. Complete demographic and baseline clinical characteristics are presented in Table 1.

**Table 1.**
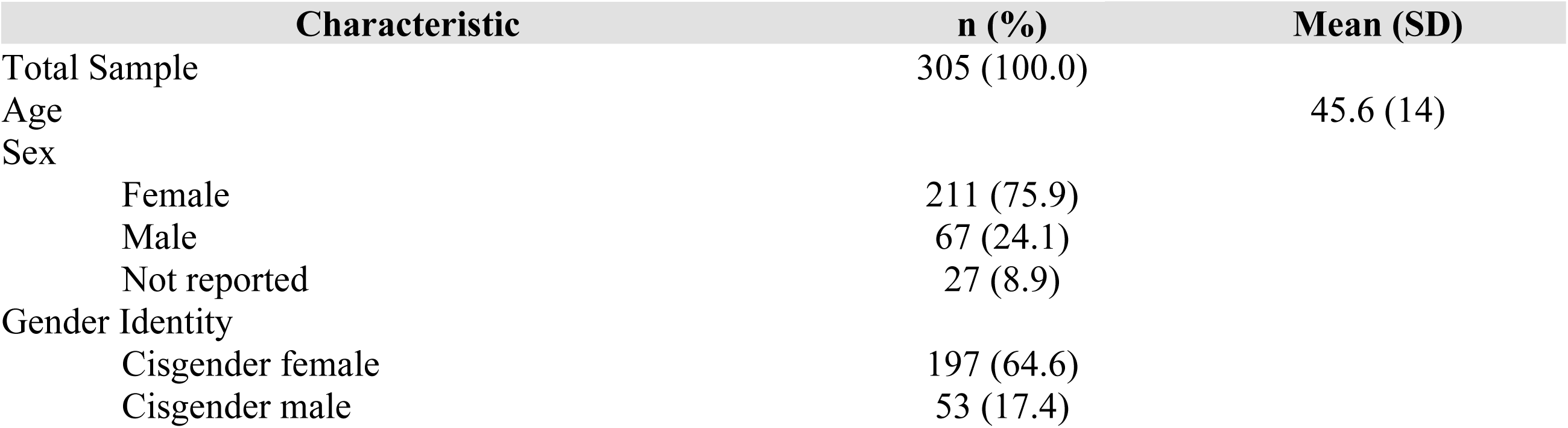
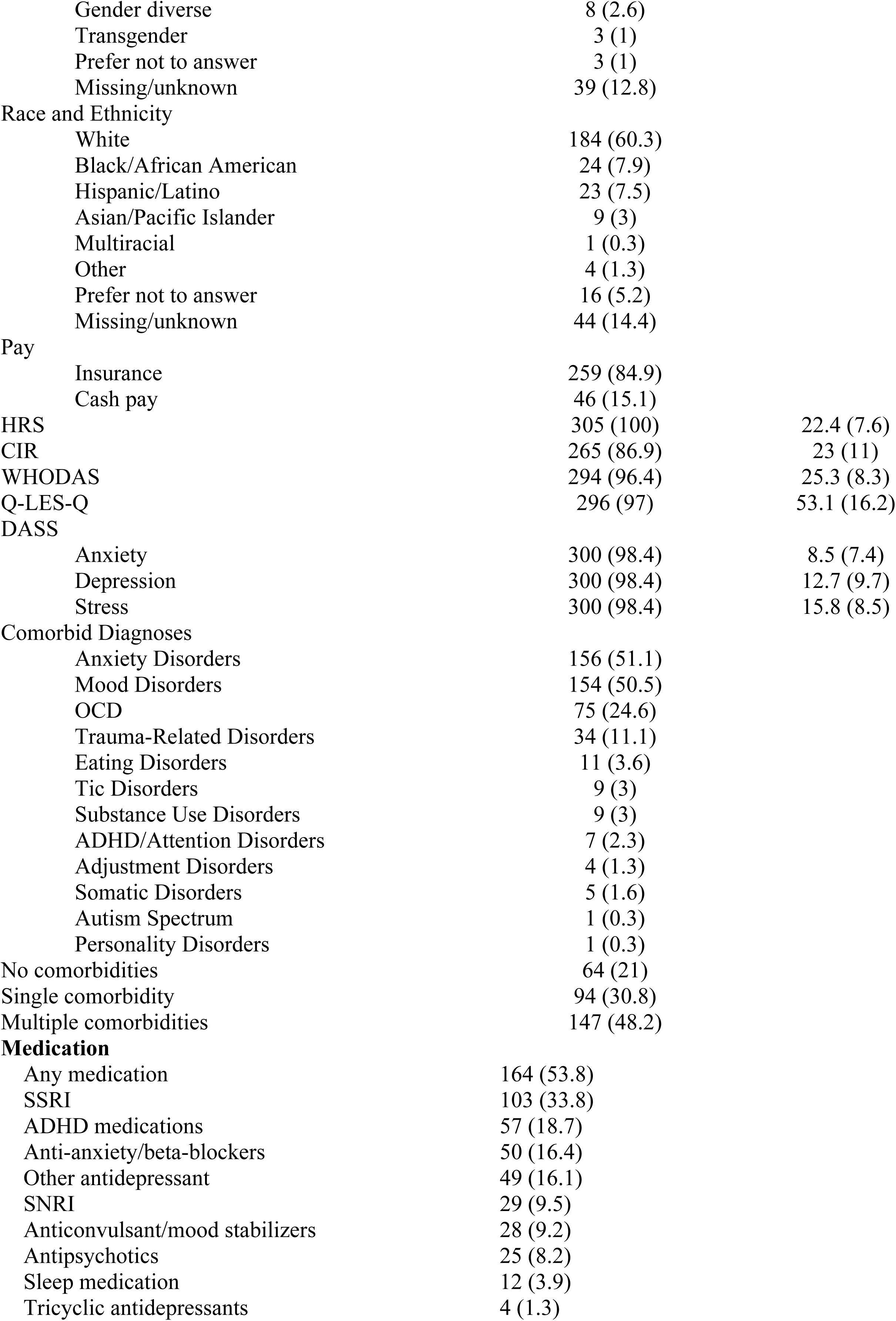
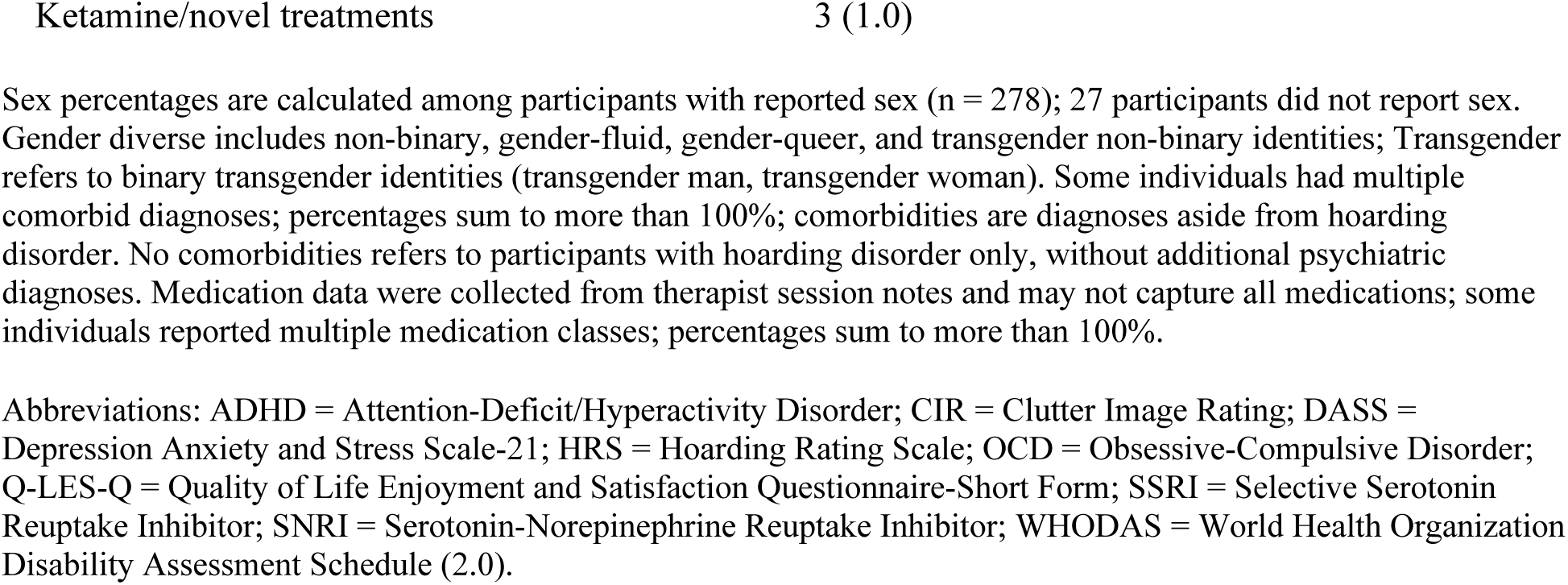
Demographic and Baseline Clinical Characteristics.

### Missing Data Analysis

Little’s MCAR test indicated that outcome data were not Missing Completely at Random, χ²(226) = 288.87, *p* = .003. Patients who completed fewer sessions were significantly younger and had lower baseline HRS severity than those who completed more sessions, while gender and race/ethnicity did not differ across completion groups; full missing data analyses are reported in the Supplementary Results. **Treatment Delivery and Engagement**

Patients completed a median of 16 hoarding-focused sessions [IQR: 8–31; *M* = 24.4, *SD* = 30.3; range: 3–352] over a median of 25.7 weeks [IQR: 14.1–41.3; *M* = 34.8, *SD* = 32.0; range: 2–190.9 weeks], at an average intensity of 0.75 sessions per week (*SD* = 0.39). Session 10 was reached by 211 patients (69.2%) and session 20 by 122 patients (40.0%). The distribution of sessions, treatment duration, and session intensity are displayed in Fig 1.

**Fig 1.**
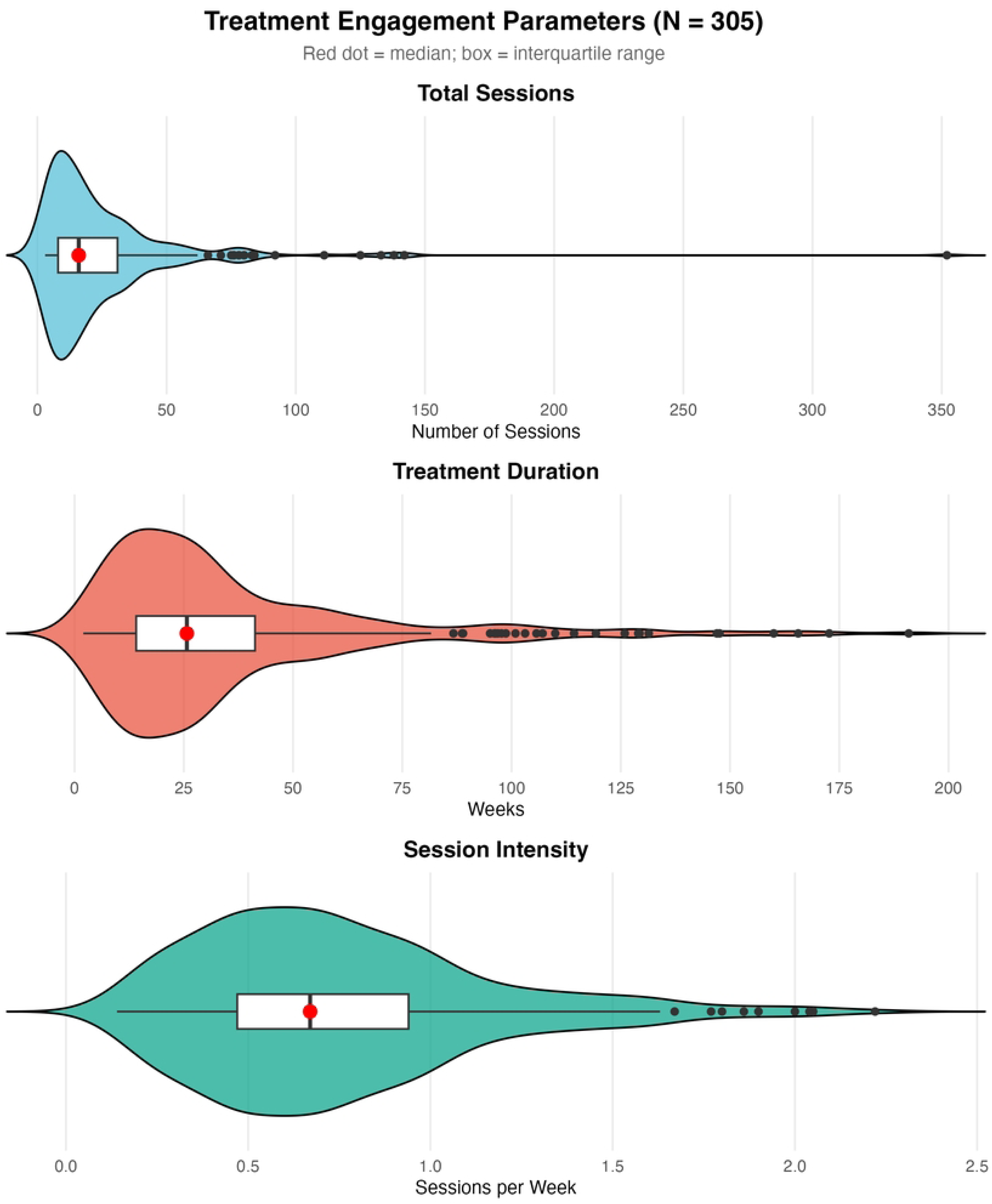
Distributions of Treatment Sessions and Duration. *Note. N* = 305. Violins represent the full distribution of values. Boxes represent the interquartile range (25th–75th percentile); horizontal lines indicate the median; red dots indicate the mean. Total sessions ranged from 3 to 352; treatment duration ranged from 2 to 190.9 weeks. The long upper tails reflect a subset of patients who engaged in extended treatment.

### Treatment Context

To characterize the broader platform treatment context, treatment history was examined across three periods: prior to, during, and following the selected hoarding episode (see S1 Fig and Supplementary Results for full details). Nearly half the sample (n = 146, 47.9%) had no prior treatment on the platform, indicating HD was their initial presenting concern. Among the 159 patients (52.1%) with prior treatment history, OCD was the predominant prior focus (n = 150, 94.3% of those with any history; median = 10 prior OCD sessions, IQR = 3–23). Concurrent OCD treatment was also common during the hoarding episode itself, with 155 patients (50.8%) having at least one OCD-coded session within their selected episode (median = 7 OCD sessions, IQR = 4–16); full within-episode session composition is reported in the Supplementary Results. Following the episode, 194 patients (63.6%) had no further recorded NOCD treatment; among the 111 who continued with NOCD treatment (36.4%), OCD was again the dominant focus (n = 99, 89.2% of continuers); full post-episode treatment details are reported in the Supplementary Results.

### Primary Outcomes: Overall Group Mean Treatment Effects

HRS scores decreased significantly across three treatment timepoints (baseline, session 10, and session 20), *F*(2, 347.46) = 53.98, *p* < .001 (see Fig 2; Table 2). HRS scores declined from 22.4 (*SD* = 7.6) at baseline to 19.8 (*SD* = 7.4) at session 10 (−2.6 points, 11.6%; Hedges’ *g* = 0.60, 95% CI [0.46, 0.75], *n* = 211) and to 18.9 (*SD* = 7.5) at session 20 (−3.5 points, 15.6%; Hedges’ *g* = 0.78, 95% CI [0.58, 0.98], *n* = 122). All pairwise comparisons were significant (all *p*s ≤ .010).

**Fig 2.**
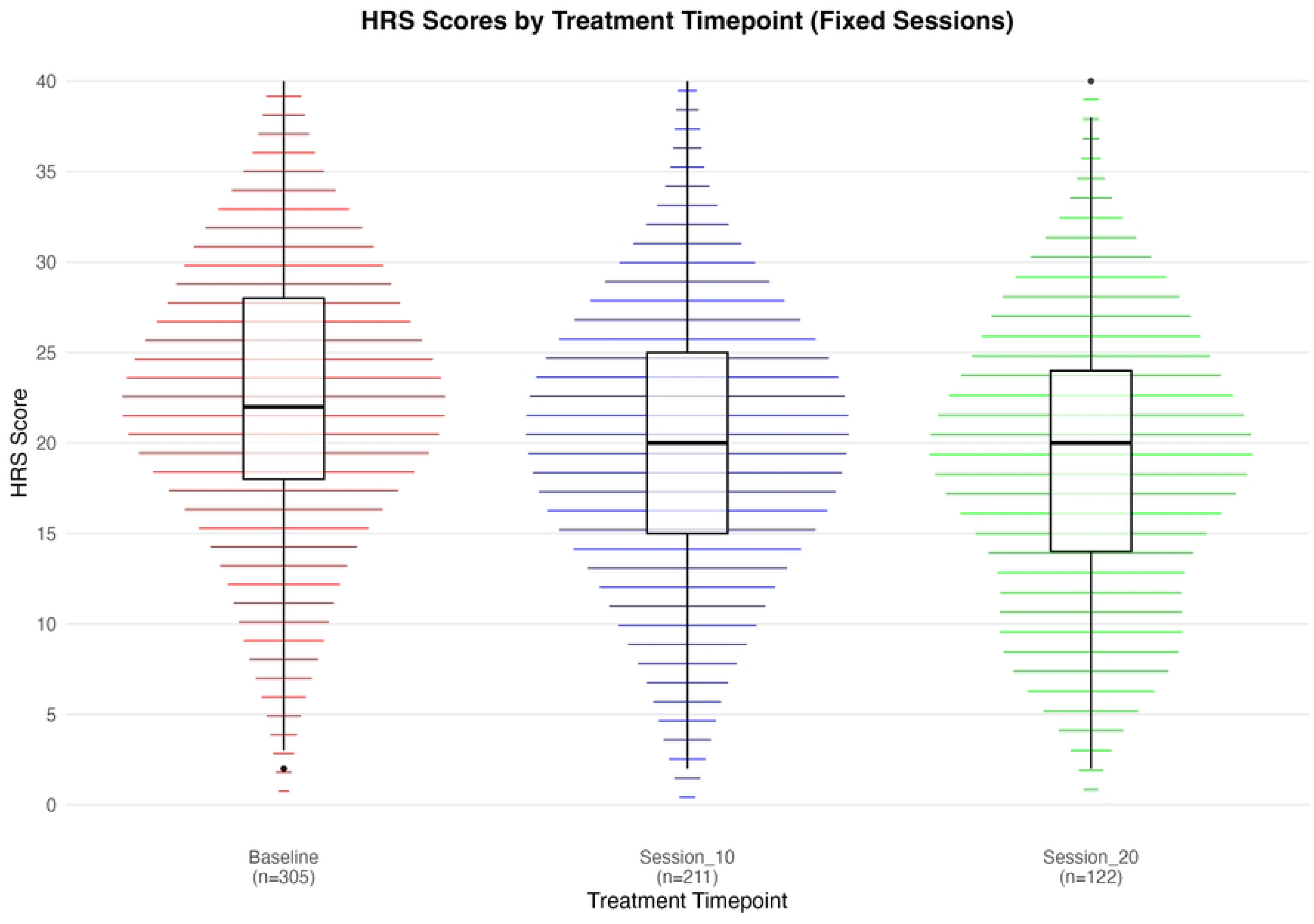
HRS Scores at Baseline, Session 10, and Session 20. Distribution of hoarding symptom severity (HRS-SR) at baseline, session 10, and session 20 for patients reaching each timepoint (baseline: *N* = 305; session 10: n = 211; session 20: n = 122). Median and interquartile ranges are indicated in the box-and-whisker plots. *p* < .001 for the effect of assessment period. Abbreviations: HRS = Hoarding Rating Scale–Self Report.

**Table 2.** Clinical Assessments by Treatment Time Point.

| Outcome | Timepoint | Valid<br>(N) | Missing<br>(N) | Mean<br>(SD) |
| --- | --- | --- | --- | --- |
| HRS | Baseline | 305<br>(100) | 0 | 22.4 (7.6) |
|  | Session 10 | 211 | 94 | 19.8 (7.4) |
|  | Session 20 | 122 | 183 | 18.9 (7.5) |
|  | Final Session | 305 | 0 | 16.4 (8.2) |
| CIR | Baseline | 265 | 40 | 23 (11) |
|  | Session 10 | 192 | 113 | 21.8 (10.5) |
|  | Session 20 | 115 | 190 | 21.7 (10) |
|  | Final Session | 288 | 17 | 19.8 (10.6) |
| DASS depression | Baseline | 300 | 5 | 12.7 (9.7) |
|  | Session 10 | 211 | 94 | 11.8 (9.2) |
|  | Session 20 | 122 | 183 | 11.3 (8.6) |
|  | Final Session | 305 | 0 | 11.1 (8.8) |
| DASS anxiety | Baseline | 300 | 5 | 8.5 (7.4) |
|  | Session 10 | 211 | 94 | 7.5 (6.6) |
|  | Session 20 | 122 | 183 | 7.4 (6.2) |
|  | Final Session | 305 | 0 | 6.9 (6.8) |
| DASS stress | Baseline | 300 | 5 | 15.8 (8.5) |
|  | Session 10 | 211 | 94 | 14.4 (8.4) |
|  | Session 20 | 122 | 183 | 14.6 (7.5) |
|  | Final Session | 305 | 0 | 13.4 (8) |
| Q-LES-Q | Baseline | 296 | 9 | 53.1 (16.2) |
|  | Session 10 | 211 | 94 | 54.3 (16.9) |
|  | Session 20 | 122 | 183 | 55 (14.5) |
|  | Final Session | 304 | 1 | 56.5 (16.2) |
| WHODAS | Baseline | 294 | 11 | 25.3 (8.3) |
|  | Session 10 | 209 | 96 | 24.9 (8) |
|  | Session 20 | 122 | 183 | 24.5 (7.3) |
|  | Final Session | 304 | 1 | 23.5 (7.9) |

Missing ns reflect patients who either discontinued treatment prior to that assessment timepoint or did not complete the assessment at that session. Patients may have discontinued due to meeting treatment goals, premature dropout, logistical reasons, or shifting clinical focus to another condition; the naturalistic design did not allow reliable differentiation between these reasons. Final session values for HRS and DASS reflect last-observation-carried-forward (LOCF) imputation; missing n=0 because all patients had at least one prior assessment for these measures. CIR, Q-LES-Q, and WHODAS were introduced partway through the study period; patients who began treatment before these measures were implemented had no prior assessment available for LOCF and are therefore missing at final session.
Abbreviations: CIR = Clutter Image Rating; DASS = Depression Anxiety and Stress Scale–21; HRS = Hoarding Rating Scale; LOCF = Last Observation Carried Forward; Q-LES-Q = Quality of Life Enjoyment and Satisfaction Questionnaire–Short Form; WHODAS = World Health Organization Disability Assessment Schedule (2.0).

### Primary Outcomes: Baseline to Final Session

From baseline to individuals’ final sessions (which varied by individual), HRS scores decreased 6.0 points from 22.4 (*SD* = 7.6) at baseline to 16.4 (*SD* = 8.2) at the final session (−26.8%; *F*(1, 304) = 200.85, *p* < .001; Hedges’ *g* = 0.81, 95% CI [0.68, 0.94], *n* = 305), with patients completing a median of 16 sessions [IQR: 8–31] (see Fig 3).

**Fig 3.**
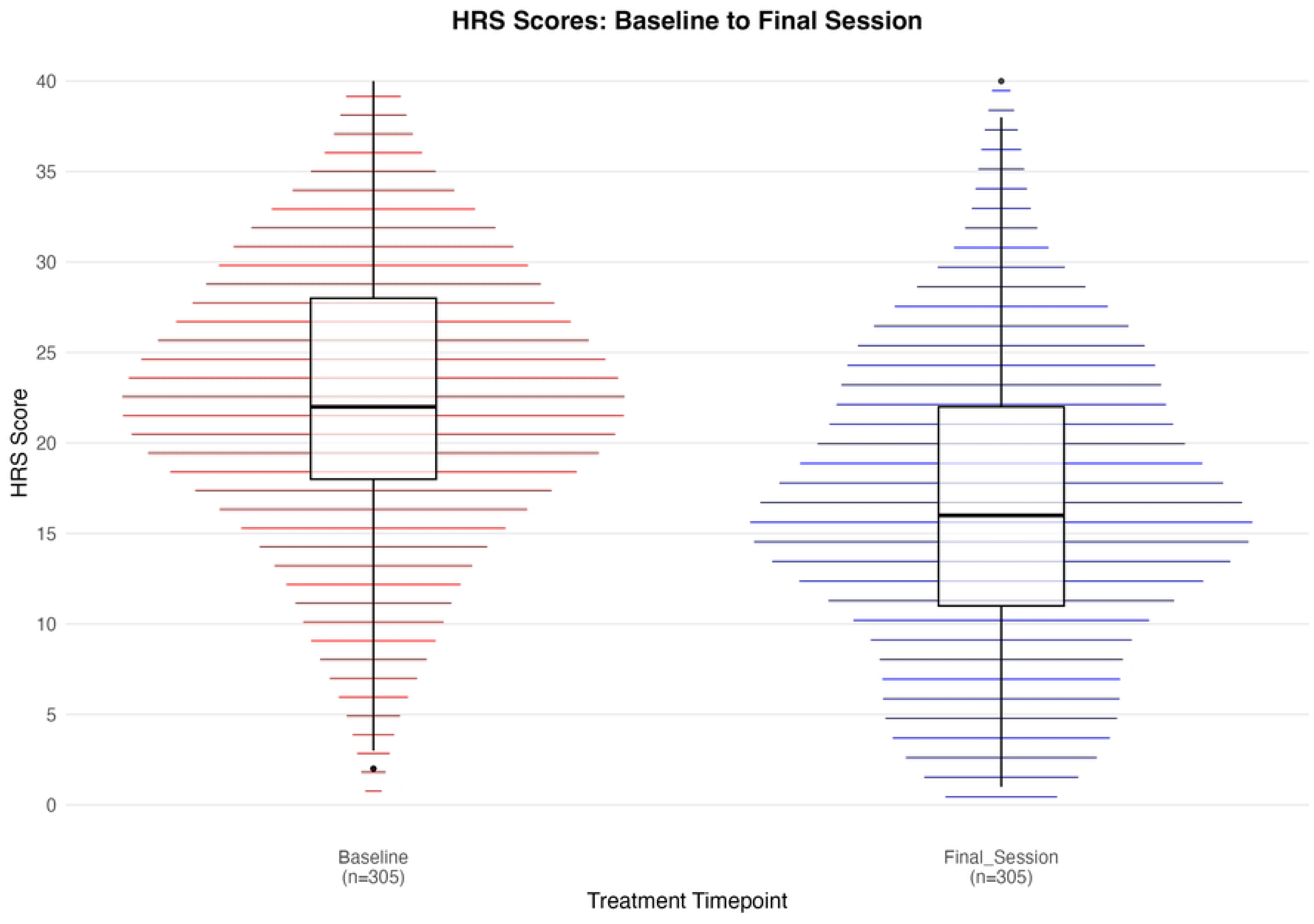
HRS Scores From Baseline to Final Session. Changes in hoarding symptom severity (HRS-SR) from baseline to final session (range: 3–352 sessions, median: 16 [IQR: 8–31]). Median and interquartile ranges are indicated in the box-and-whisker plots. *p* < .001 for the effect of assessment period.

### Individual-Level Treatment Response

Median percent improvement in HRS scores was 13.0% [IQR: 0–31.6%] at session 10, 24.6% [IQR: 4.6–37.2%] at session 20, and 25.0% [IQR: 3.0–46.7%] at the final session. At session 10, 33.2% (70/211) achieved ≥25% reduction, 19.4% (41/211) achieved ≥35% reduction, 14.2% (30/211) achieved ≥45% reduction, and 8.5% (18/211) achieved ≥55% reduction; 22.7% (48/211) showed any worsening. At session 20, 50.0% (61/122) achieved ≥25% reduction, 29.5% (36/122) achieved ≥35% reduction, 20.5% (25/122) achieved ≥45% reduction, and 9.8% (12/122) achieved ≥55% reduction; 19.7% (24/122) showed any worsening. At the final session, 51.8% (158/305) achieved ≥25% reduction, 39.3% (120/305) achieved ≥35% reduction, 26.2% (80/305) achieved ≥45% reduction, and 18.0% (55/305) achieved ≥55% reduction; 19.7% (60/305) showed any worsening. The majority with apparent worsening reflected small absolute fluctuations (median 3.0 points), with only 3 patients (1.0%) exceeding the threshold for reliable deterioration on the RCI. Among the 266 patients who began treatment above the clinical cutoff (HRS ≥ 14), 16.2% (32/198) achieved remission by session 10, 20.9% (24/115) by session 20, and 27.4% (73/266) by the final session.

### Reliable Change Index

Reliable change analyses indicated that an RCI threshold of 8.15 HRS points was required to exceed measurement error in the present sample (*SD* = 7.59; ICC = 0.85; 40). From baseline to final session (N = 305), 36.4% of patients (n = 111) demonstrated reliable improvement, 62.6% (n = 191) showed no reliable change, and 1.0% (n = 3) demonstrated reliable deterioration. Among the 266 patients who began treatment above the clinical cutoff (HRS ≥ 14), 22.9% (n = 61) achieved clinically significant change, defined as reliable improvement combined with a final session score below the clinical threshold (HRS < 14). Session 20 RCI analyses and concordance between RCI and percentage-based thresholds are reported in the Supplementary Results.

### Time to Response

Among the 120 patients (39.3%) who achieved and maintained ≥35% HRS reduction through their final session (i.e., whose final observed HRS assessment also met the ≥35% reduction threshold), median time to first response was session 9 (IQR: 5–19), with 54.2% responding within the first 10 sessions, 77.5% within the first 20 sessions, and 22.5% requiring more than 20 sessions to first respond. Among the 185 patients who either never achieved ≥35% HRS reduction or did not sustain it through their final session, mean total sessions was 20.8 and mean baseline HRS was 21.6, suggesting that non-responders had slightly lower baseline severity and engaged in somewhat shorter treatment courses. Of the 161 patients (52.8%) who ever achieved ≥35% HRS reduction at any point, 120 (74.5%) maintained that response through their final session. A secondary time-to-response analysis using the more stringent Jacobson-Truax clinically significant change criterion (reliable improvement on the RCI combined with crossing from clinical to sub-clinical range; n = 61 among patients clinical at baseline) yielded broadly consistent findings: median time to first response was session 11 (IQR: 8–19), with 47.5% responding within 10 sessions, 78.7% within 20 sessions, and 21.3% requiring more than 20 sessions. Full results of the secondary analysis are reported in the Supplementary Results.

### Depression, Anxiety, Stress, Quality of Life, and Disability

There were significant improvements across all secondary outcomes from baseline to final session (see Table 2). Anxiety symptoms (DASS-21 Anxiety) decreased from 8.5 (*SD* = 7.4) to 6.9 (*SD* = 6.9), an 18.6% reduction, *F*(1, 299) = 22.36, *p* < .001; Hedges’ *g* = 0.27, 95% CI [0.16, 0.39]. Depression symptoms (DASS-21 Depression) decreased from 12.7 (*SD* = 9.7) to 11.1 (*SD* = 8.8), a 12.6% reduction, *F*(1, 299) = 13.18, *p* < .001; Hedges’ *g* = 0.21, 95% CI [0.09, 0.32]. Stress (DASS-21 Stress) decreased from 15.8 (*SD* = 8.5) to 13.4 (*SD* = 8.0), a 14.8% reduction, *F*(1, 299) = 27.47, *p* < .001; Hedges’ *g* = 0.30, 95% CI [0.19, 0.42]. Clutter severity (CIR) decreased from 23.0 (*SD* = 11.0) to 19.6 (*SD* = 10.5), a 14.7% reduction, *F*(1, 264) = 58.18, *p* < .001; Hedges’ *g* = 0.47, 95% CI [0.34, 0.59]. Functional disability (WHODAS) decreased from 25.3 (*SD* = 8.3) to 23.5 (*SD* = 7.9), a 6.9% reduction, *F*(1, 293) = 28.60, *p* < .001; Hedges’ *g* = 0.31, 95% CI [0.19, 0.43]. Quality of life (Q-LES-Q) increased from 53.1 (*SD* = 16.2) to 56.5 (*SD* = 16.2), a 6.8% improvement, *F*(1, 295) = 20.08, *p* < .001; Hedges’ *g* = 0.26, 95% CI [0.14, 0.38]. All outcomes remained significant after Benjamini-Hochberg correction for multiple comparisons (all adjusted *p*s < .001).

### Moderators of Treatment Response

Baseline HRS severity significantly moderated treatment response such that patients with greater baseline severity showed larger absolute improvements, ranging from 1.2 points at mild baseline severity (HRS = 10) to 9.0 points at severe baseline severity (HRS = 30). Both total sessions and treatment duration significantly moderated outcomes such that more sessions and longer duration were associated with larger HRS reductions, consistent with a dose-response pattern. Age marginally moderated treatment response, with older patients showing slightly greater improvement. Comorbidity status and sex did not significantly moderate outcomes.

To examine whether prior OCD treatment experience facilitated HD treatment outcomes, patients with ≥3 prior OCD sessions before their HD episode (n = 115, 37.7%) were compared to those without (n = 190, 62.3%). Analyses found that patients with prior OCD treatment did not differ significantly from those without (n = 190) in HRS change after controlling for baseline HRS severity, β = -0.98, *p* = .253, or in percentage symptom reduction, *p* = .379, or response rates, χ²(1) = 0.82, *p* = .365, despite having lower baseline HRS severity (*M* = 18.9 vs *M* = 24.5). Full moderator and prior OCD treatment statistics are reported in the Supplementary Results.

## Discussion

The present study examined real-world treatment outcomes for adults with HD receiving therapist-delivered video CBT through a specialty online therapy platform. This represents the largest naturalistic investigation of therapist-delivered video CBT for HD to date. Across 305 adults, hoarding symptom severity declined significantly from baseline to final session, with a mean reduction of 26.8% and a large effect size (g = 0.81), broadly consistent with meta-analytic benchmarks from controlled trials of in-person CBT (15,16). Notably, the mean final HRS score of 16.4 remained above the clinical cutoff of 14, indicating that at the end of treatment, the average patient still had hoarding symptoms within the clinical range, a pattern observed consistently across prior HD trials (14). At the individual patient level, 39.3% achieved ≥35% HRS reduction and 27.4% of those who began treatment above the clinical threshold achieved remission. Treatment additionally produced significant improvements across all secondary outcomes. These findings demonstrate that therapist-delivered video CBT for HD is feasible at scale and produces outcomes consistent with controlled research settings. This has important implications for expanding access to care for a condition historically underserved by mental health systems.

Treatment gains followed a progressive trajectory, with modest early improvements at session 10 (11.6%) accelerating through session 20 (15.6%) and continuing to the final session (26.8%), consistent with HD requiring sustained engagement to produce meaningful gains (14,15,23). The final effect size (g = 0.81) aligns with meta-analytic estimates from controlled in-person trials (g = 0.82–1.11; 15,16,18), suggesting that therapeutic gains observed in research settings generalize to real-world video therapy delivery, extending prior evidence from much smaller pilot telehealth trials (n = 7–10; 26,27).

However, direct comparison of individual-level response rates with prior HD trials is complicated by heterogeneity in how clinically significant change has been operationalized across studies. The Tolin et al. [15] meta-analysis reported clinically significant change rates of 24–43% using Jacobson-Truax methods on the SI-R, a different measure with different psychometric properties; thus, cross-measure comparisons should be interpreted cautiously. In the present study, 39.3% of patients achieved ≥35% HRS reduction by the final session, with 26.2% achieving ≥45% and 18.0% achieving ≥55% reduction.

Among the patients who began treatment above the clinical threshold, 27.4% achieved remission, crossing into the sub-clinical range during treatment. These findings suggest that a meaningful minority achieve substantial gains, while the majority show partial or no meaningful symptom reduction; notably, 62.6% of patients showed no reliable change on the RCI, underscoring that most patients did not achieve clinically significant gains. This suggests that there is a continued need for treatment innovation in HD, as previously emphasized in the broader literature (15,23).

Reliable change analyses provided a more stringent, measurement-error-adjusted picture of individual-level change. An RCI threshold of 8.15 points was required to exceed measurement error (SD = 7.59; ICC = 0.85; 40); 36.4% of patients met this threshold at the final session, and 22.9% of eligible patients achieved clinically significant change (reliable improvement combined with a final session score below the clinical cutoff), consistent with patient-level rates in prior controlled trials (15,43). Reliable deterioration was rare (1.0% at final session), and of the 60 patients showing any HRS increase at the final session, only 3 exceeded the reliable deterioration threshold, indicating that apparent worsening largely reflected small measurement fluctuations. RCI analyses also demonstrated strong concordance with percentage-based thresholds, with convergence highest at ≥35% and ≥55% reductions and lowest at ≥25%, consistent with ≥35% representing a clinically meaningful threshold on the HRS-SR, though direct validation against independent benchmarks is warranted (see Supplementary Results for full concordance analyses).

Among patients who achieved and maintained ≥35% HRS reduction (39.3%), median time to first response was session 9, with 54.2% responding within the first 10 sessions and 77.5% within the first 20 sessions. To our knowledge, session-level time-to-response data have not been previously reported for HD. Prior HD trials have generally reported pre-post outcomes without examining session-level response trajectories (12,15), making direct comparison with the present findings difficult. Nevertheless, the present finding that over half of eventual responders showed meaningful symptom reduction within 10 sessions is clinically informative, as CBT for hoarding is traditionally characterized as requiring a high number of sessions, and indeed this was the case for many patients in the current study, as 22.5% of responders required more than 20 sessions to first respond. The findings suggest there may be a substantial subgroup who respond relatively early in treatment. Whether early response serves as a prognostic signal in HD, as it does in CBT for OCD (44,45), and what patient and/or therapist features could predict early response warrants prospective investigation. Further, 74.5% of those who ever achieved ≥35% reduction maintained it through their final session, underscoring that response, once achieved, is generally durable.

Treatment produced significant, although smaller magnitude, improvements across all secondary outcomes (g = 0.21–0.47), consistent with controlled trials where gains in functioning and mood are typically smaller than improvements in core hoarding symptoms (15,16). Clutter severity showed a 14.7% reduction, aligning with prior evidence identifying clutter as the most treatment-resistant HD domain (15). Depression, anxiety, and stress improvements were modest, consistent with meta-analytic findings (g ≈ 0.45 for depression; 16) and observations that mood symptoms show limited improvement relative to hoarding-specific gains (12). Quality of life and disability are rarely assessed as primary HD outcomes (23,46) and everyday functioning typically does not normalize following HD treatment even when symptom reduction is achieved (15). Significant improvements in both domains, though modest, extend the evidence base on functional recovery in HD.

Greater baseline severity predicted larger absolute HRS reductions, consistent with prior evidence that baseline severity does not preclude improvement and may predict greater absolute change (12,15,17). Although regression to the mean could theoretically contribute to this pattern, naturalistic and longitudinal data indicate that hoarding symptoms are highly stable without treatment, showing little spontaneous remission and gradual worsening over time (16,47,48), suggesting that observed improvements in higher-severity patients are unlikely to be fully explained by measurement regression.

Further, a dose-response pattern emerged, with more sessions and longer duration associated with larger reductions, though this finding warrants caution: patients who engaged longer also had higher baseline severity, and the missing data analysis confirmed that earlier discontinuation was associated with lower baseline HRS, introducing the possibility that selection effects partially explain this pattern. Age marginally moderated response, with older patients showing slightly greater improvement, diverging from meta-analytic findings associating younger age with better outcomes (15). Neither comorbidity status nor sex significantly moderated outcomes, suggesting that patients with co-occurring mood and anxiety disorders responded comparably to those without, consistent with prior evidence that comorbidity burden alone does not reliably attenuate response to HD-focused CBT (49). The treatment history data reveal an important contextual feature of the present sample, all of whom accessed HD treatment through an OCD-spectrum specialty platform: 52.1% received OCD treatment prior to HD treatment and 50.8% receiving both OCD and HD sessions. This level of OCD co-occurrence substantially exceeds the ∼20% rate in community-based HD samples (50,51) and likely reflects the platform’s specialization. Research suggests that comorbid OCD is a key driver of treatment-seeking in HD (52), and the present sample likely represents patients who sought care primarily for OCD and subsequently engaged in HD-focused treatment, rather than a population presenting with HD as their primary concern without prior OCD treatment. This limits generalizability to the broader HD population, which includes many individuals who present through housing, social care, or crisis pathways rather than mental health services (19,20).

To examine whether prior OCD treatment facilitated HD outcomes, given that ERP skills might transfer to HD-focused CBT, patients with ≥3 prior OCD sessions (n = 115, 37.7%) were compared to those without. After controlling for baseline severity differences between groups, prior OCD treatment did not significantly moderate HD outcomes, suggesting that patients engaging with HD treatment after prior OCD treatment respond comparably to those presenting with HD as their initial concern.

The present findings contribute to a growing literature supporting therapist-delivered video CBT for HD. Conducting sessions via video within patients’ own homes addresses a distinctive feature of HD treatment: the living environment is both the site of symptoms and the most ecologically valid context for sorting, discarding, and behaviour change (21,24). Between-session platform features, including therapist messaging and peer community access, may have further sustained motivation and homework engagement (29,53). Notably, 47.9% of patients had no prior NOCD treatment history, indicating that HD represented their initial presenting concern on the platform. While prior care received outside NOCD cannot be determined from these data, this finding suggests telehealth may reduce barriers of cost, geography, and reluctance to allow others into the home that are characteristic of this population (21,24).

### Limitations

Several limitations warrant consideration. The retrospective naturalistic design without a control group precludes causal inferences and cannot rule out natural symptom fluctuations (and, related to this, regression to the mean), or non-specific therapeutic factors. Treatment fidelity was not formally assessed. The use of LOCF for missing final assessments may additionally introduce bias by assuming stability in scores between observations. Although missing data analyses identified baseline predictors of earlier discontinuation, we cannot rule out that patients who discontinued earlier had different outcomes than those who remained engaged, whether due to symptom improvement, premature discontinuation due to treatment worsening or logistical reasons such as loss of health insurance, or transition to other treatment. Future research of therapist-delivered video CBT for hoarding disorder, including randomized controlled trials with active treatment controls, therapist fidelity assessments, and long-term follow-up, will provide additional important information about this treatment.

The HRS-SR relies solely on self-report, used in a population known for limited insight into symptom severity (14). In addition, there is not a validated percentage-based response threshold for this measure, necessitating the proxy use of a benchmark approximated from the SI-R literature; response classifications should be interpreted accordingly. Although prior OCD treatment history did not significantly moderate HD treatment outcomes in the present analyses, other factors among the 52.1% of patients with prior OCD treatment, including familiarity with CBT approaches, prior experience navigating therapy, and existing treatment momentum, may have facilitated engagement in ways not fully captured by symptom change alone. Inclusion criteria requiring valid baseline and follow-up HRS assessments mean the analytic sample represents patients who engaged sufficiently to complete repeated assessments and may not be representative of all treatment-seeking individuals with HD.

## Conclusions

This study provides the largest naturalistic examination of therapist-delivered video CBT for HD to date, demonstrating that outcomes consistent with controlled trials are achievable in real-world delivery. This format can reach patients who have not previously accessed care, helping fill the substantial evidence-based health care accessibility gap for HD.

## Methods

### Sample

This retrospective, observational analysis examined clinical data from adults who received treatment for HD through an online specialty therapy platform between September 2021 and February 2026. Patients were identified through ICD-10 diagnostic codes (F42.3 for HD) assigned as primary or secondary diagnosis by trained therapists. Hoarding-focused sessions were defined as sessions where HD was coded as the diagnosis being treated. Data were accessed for research purposes in February 2026. Authors did not have access to information that could identify individual participants; all data were de-identified prior to analysis.

Inclusion criteria required: (1) age ≥18 years; (2) ≥3 hoarding-focused sessions within a treatment episode; (3) ≥2 HRS assessments within the episode (n = 186 excluded); and (4) valid baseline HRS assessment completed within the first two sessions (n = 96 excluded). The final analytic sample included 305 adults (M_age=_ 45.6 years, SD_age_ = 14.0; 75.9% female; 60.3% White). Complete demographic and baseline characteristics are presented in Table 1. Details regarding treatment episode selection are provided in the Supplementary Materials.

### Materials

Clinical assessments included the Hoarding Rating Scale-Self Report (HRS-SR; 31) for hoarding symptom severity, the Clutter Image Rating (CIR; 32) for visual assessment of clutter, and the Diagnostic Interview for Anxiety, Mood, and Obsessive-Compulsive and Related Neuropsychiatric Disorders (DIAMOND; 33) for diagnostic evaluation of HD and comorbidities. Secondary measures assessed depression, anxiety, and stress using the Depression, Anxiety, and Stress Scale (DASS-21; 34), quality of life using the Quality of Life Enjoyment and Satisfaction Questionnaire-Short Form (Q-LES-Q-SF; 35), and disability and functioning using the World Health Organization Disability Assessment Schedule 2.0 (WHODAS 2.0; 36). Detailed descriptions of all secondary measures are provided in the Supplementary Materials.

#### Hoarding Rating Scale-Self Report (HRS-SR)

The HRS-SR is a 5-item self-report measure of hoarding symptom severity with scores ranging from 0 to 40; a score of ≥14 indicates clinically significant hoarding (31,37). Full psychometric details are provided in the Supplementary Materials.

### Procedures

#### Therapist Training and Treatment Delivery

Therapists who provided treatment completed NOCD’s onboarding training in OCD and Related Disorders. This involves a mix of live and video didactic training. NOCD’s Hoarding training protocol (an additional five hours of hoarding didactics and attendance to a weekly case consultation group for hoarding) is open for clinicians who are interested in treating HD. Training primarily focused on the following areas: (1) the nature and assessment of HD, (2) conceptualization of hoarding with emphasis on maintaining factors, (3) patient-focused psychoeducation, (4) treatment planning, and (5) completion of treatment activities within and between therapy sessions. Upon completion of training, therapists were eligible to begin providing treatment to patients. Therapists were required to attend weekly group case consultation meetings focused on HD treatment. Therapists were trained to recommend two 60-minute sessions per week at a minimum until achieving clinically significant symptom reduction. Treatment incorporated cognitive-behavioral approaches for hoarding, including motivational enhancement, psychoeducation, sorting/discarding practice, and reducing acquisition. Much of the decluttering involved the use of a four box method approach, where the Member received four equal sized boxes for items to keep, donate, recycle, and discard. Once all four boxes were filled, a trusted friend or family member took three of the four boxes (the Keep box was kept at the home) and did as instructed with them (donate, etc.) All sessions were conducted via Zoom (US Health Insurance Portability and Accountability Act–compliant version). Patients could join sessions from any computer or personal electronic device with internet access. Therapists and patients were required to be on video throughout all sessions. Patients also had access to in-app messaging with their therapist for support between sessions and an online community platform.

### Data Processing

Baseline assessments were defined as the first HRS assessment completed within the first two hoarding-focused sessions of the selected treatment episode. Outcomes were analyzed at sessions 10 and 20 to capture early and mid-treatment response, as well as each patient’s final treatment session. These timepoints were selected to reflect typical hoarding treatment protocols, which generally range from 16–26 sessions (12,15). When assessments were missed, a last-observation-carried-forward (LOCF) approach was applied. Detailed data processing procedures are described in the Supplementary Materials.

### Data Analysis

Treatment parameters, including total hoarding-focused sessions, duration, intensity (sessions per week), and session composition, were calculated for each patient. Little’s MCAR test (38) was used to evaluate missing data assumptions; baseline characteristics were compared across treatment completion groups to characterize predictors of missingness. Full missing data analyses are reported in the Supplementary Results.

Primary and secondary outcomes were analyzed using linear mixed models with treatment timepoint (baseline, session 10, session 20, final session) as a fixed factor and patient as a random factor, utilizing restricted maximum likelihood estimation. Individual-level changes were calculated by comparing baseline with each follow-up assessment.

Individual-level treatment response was examined across a range of percentage reduction thresholds (≥55%, ≥45%, ≥35%, ≥25%, >0%, and worsening), given the absence of a validated response threshold for the HRS-SR in the published literature. Individual-level reliable change was examined using the Reliable Change Index (39), which determines whether change in HRS score from baseline to each timepoint exceeded that expected by measurement error alone. RCI was calculated using the test-retest reliability coefficient reported for the Hoarding Rating Scale-Interview (ICC = 0.85; 40), as no equivalent coefficient has been published for the English internet-administered HRS-SR. Clinically significant change was defined as the combination of reliable improvement on the RCI and a final session HRS score below the established clinical cutoff (HRS < 14), consistent with the Jacobson-Truax two-step criterion (39). Only patients who began treatment above the clinical threshold (HRS ≥ 14; n = 266) were eligible for this classification. Remission was defined as a final session HRS score below the established clinical cutoff (HRS < 14; 31,37), reflecting movement from the clinical to the sub-clinical range; remission rates are reported only among patients who began treatment above this threshold (n = 266). Time-to-response was defined as the first session at which a patient achieved ≥35% HRS reduction from baseline; analyses were restricted to patients who achieved and maintained this threshold through their final session (n = 120, 39.3%). This threshold was selected as the closest available benchmark, corresponding to the percentage reduction implied by the only standardised clinically significant change criterion in the HD literature, a ≥20-point Saving Inventory Revised (SI-R) (41) reduction, which, at typical untreated HD baseline scores, corresponds to approximately 35% symptom reduction (23,42). A secondary time-to-response analysis was conducted using the Jacobson-Truax clinically significant change criterion (reliable improvement on the RCI combined with a final session HRS score below the clinical cutoff; restricted to patients with baseline HRS ≥ 14, n = 266); results are reported in the Supplementary Results.

Effect sizes were computed using Hedges’ *g* with 95% confidence intervals. Benjamini-Hochberg correction was applied to the seven primary and secondary outcome tests; moderation analyses were considered exploratory and are reported without correction. All analyses were conducted in R (version 4.3.1).

Given the high prevalence of prior OCD treatment in the sample, a supplementary analysis examined whether prior OCD treatment (defined as ≥3 OCD-coded sessions before the HD episode) moderated HD treatment outcomes, using ANCOVA with baseline HRS as a covariate.

### Ethical Considerations

These analyses did not require research ethics board review, as they do not meet the definition of Human Subjects Research under 45 CFR 46.102(e). This was a secondary analysis of de-identified clinical data obtained retrospectively, with all procedures compliant with NOCD’s privacy policy and applicable data protection laws. As this study used de-identified, retrospectively collected clinical data and did not involve direct participant contact, individual informed consent was not required.

### Patient and Public Involvement

Patients were not directly involved in the design, conduct, reporting, or dissemination plans of this research. This was a retrospective analysis of de-identified clinical data. Patient perspectives are indirectly represented through the naturalistic treatment outcomes analysed, which reflect real-world clinical care delivered through the NOCD platform.

## Data Availability

The data underlying this study are proprietary clinical data held by NOCD, Inc. and cannot be made publicly available due to patient privacy protections and commercial confidentiality restrictions. The data contain sensitive mental health information collected in the context of clinical care, and public release is not permitted under NOCD's privacy policy and applicable data protection laws. Aggregate summary statistics supporting the findings are available upon reasonable request to the corresponding author at.

## Data Availability Statement

The data in this study are proprietary clinical data held by NOCD, Inc. and are not publicly available due to patient privacy and commercial confidentiality restrictions. Reasonable requests for aggregate summary statistics may be directed to the corresponding author.

## Acknowledgments

The authors thank the NOCD therapists and Member Advocates for their help in facilitating treatment and care experiences.

## Author Contributions

JDF: Conceptualization, Investigation, Methodology, Project Administration, Supervision, Writing – Original Draft, Writing – Review & Editing.

CCB: Conceptualization, Investigation, Methodology, Writing – Original Draft, Writing – Review & Editing.

PBM: Supervision, Writing – Original Draft, Writing – Review & Editing.

NRF: Supervision, Writing – Original Draft, Writing – Review & Editing.

MN: Supervision, Writing – Original Draft, Writing – Review & Editing.

NL: Data Curation, Writing – Review & Editing.

LT: Project Administration, Supervision, Resources, Writing – Review & Editing.

SMS: Conceptualization, Supervision, Resources, Writing – Review & Editing.

AR: Data Curation, Formal Analysis, Investigation, Methodology, Visualization, Writing – Original Draft, Writing – Review & Editing.

